# Application of the J-CTO Score to Intraplaque Guidewire Tracking-Based Recanalization for In-Stent Chronic Total Occlusions

**DOI:** 10.1101/2024.08.21.24312395

**Authors:** Chieh-Yu Chen, Chi-Hung Huang, Jen-Fang Cheng, Chien-Lin Lee, Jiun-Yang Chiang, Shih-Chi Liu, Chi-Jen Chang, Chia-Pin Lin, Cheng-Ting Tsai, Jun-Ting Liou, Chia-Ti Tsai, Yi-Chih Wang, Juey-Jen Hwang

## Abstract

**Background:** The application of the J-CTO score for in-stent chronic total occlusion (CTO) recanalization remains unclear. We aimed to compare the role of J-CTO score in in-stent and de novo CTO interventions using intraplaque guidewire tracking techniques.

**Methods:** The application of the J-CTO score to assess procedural feasibility and guidewire crossing time for in-stent (N=74, 14.6%) and de novo CTO (N=434, 85.4%) interventions was evaluated in consecutive 508 patients (64.1±11.6 years, 446 men). Failed intraplaque tracking (N=3) or guidewires crossing (N=35) was considered procedural failures (38/508=7.5%).

**Results:** The procedural success rate for de novo CTOs significantly declined when the J-CTO score was >2 (85 vs. ≤2: 97%, p<0.001), but was comparable for in-stent CTOs (>2: 96 vs. ≤2: 100%, p=0.400). Among 470 patients with successful recanalization, the guidewire crossing time ≥30 minutes was required less for in-stent than for de novo CTOs (OR=0.40, 95% CI=0.18-0.86) with J-CTO score ≥2 in multivariate analysis. For those with successful antegrade-only wiring, the guidewire crossing time shown by Kaplan–Meier curves was significantly related to the J-CTO score for either in-stent (N=72) or de novo (N=370) CTOs (both p<0.001 by log-rank test). However, only blunt stump (15.0±5.6 min) and occlusion ≥20mm (16.2±5.6 min) were independent time-determining factors of guidewire crossing (both p<0.01) for in-stent CTOs.

**Conclusion:** With the intraplaque tracking strategy, the effects of the J-CTO score on procedural feasibility and guidewire crossing time differ for in-stent and de novo CTOs. Therefore, the J-CTO score should be cautiously interpreted during in-stent CTO interventions.

## INTRODUCTION

Recanalization of chronic total occlusion (CTO) remains a challenging procedure for treating coronary artery disease, and accounts for approximately 15% of all percutaneous coronary interventions (PCI).^1,2^ Among CTO-PCIs, in-stent occlusion is a distinct category, and its incidence rate is approximately 15% in a pooled data of four multicenter registries.^3^ For CTO-PCI, the Japan CTO (Multicenter Chronic Total Occlusion Registry of Japan: J-CTO) score was initially developed to grade the difficulty of crossing the occluded lesion within 30 minutes,^4^ and later to evaluate the feasibility of procedural success.^5^ The score has been subsequently applied to nearly all CTO-PCI studies including those comparing procedural features between in-stent and de novo CTO-PCIs.^6–10^ However, the parameters of the J-CTO score were initially derived from native lesions,^4^ and the application of the scoring system to in-stent occlusions remains unclear.

Furthermore, the comparable technique success rates between in-stent and de novo CTO-PCIs in the literature are based upon substantially diverse recanalization techniques or devices used for each group.^3^ For example, a preponderance of antegrade wiring techniques for in-stent CTO and retrograde techniques for de novo CTO is particularly evident. For CTO interventionists, the easily visible stent contour significantly improves the accessibility of guidewire tracking through the vessel course of the CTO segment. Therefore, the rationale of wiring-based intraplaque crossing techniques is more distinct for in-stent than for de novo CTO-PCI. As the application of the J-CTO score to in-stent CTO-PCI with intraplaque guidewire tracking techniques remains unclear, this study aimed to compare it with de novo CTO-PCI in terms of procedural feasibility and guidewire crossing time.

## METHODS

### Study Design and Population

This prospective registry study was initiated and approved by the Ethics Committee of National Taiwan University Hospital (201904023RINC) in 2019. Retrospective data collection beginning in August 2014 was approved (201907064RIND). Based on the concept of intraplaque guidewire tracking for CTO-PCI developed by the Taiwan True-Lumen Tracking club,^11^ the period of retrospective data collection was determined by the introduction of the Gaia series (Asahi Intecc Medical, Japan) guidewires in Taiwan. The Gaia series guidewire was designed specifically for intraplaque tracking. Written informed consent was obtained from all the participants. From August 2014 to March 2023, 508 consecutive patients (64.1±11.6 years, 446 men) with CTO-PCI using intraplaque guidewire tracking strategy were enrolled for analyses (Figure 1). In-stent CTO was defined as occlusion located within a previously deployed stent or within 5 mm proximal and distal to it.^12^ The presence of chronic kidney disease (CKD) was defined as an estimated glomerular filtration rate of <60 ml/min/1.73 m^2^. Patients with congestive heart failure must meet the Framingham criteria with a left ventricular ejection fraction <50% before the index procedure.

**Figure 1.**
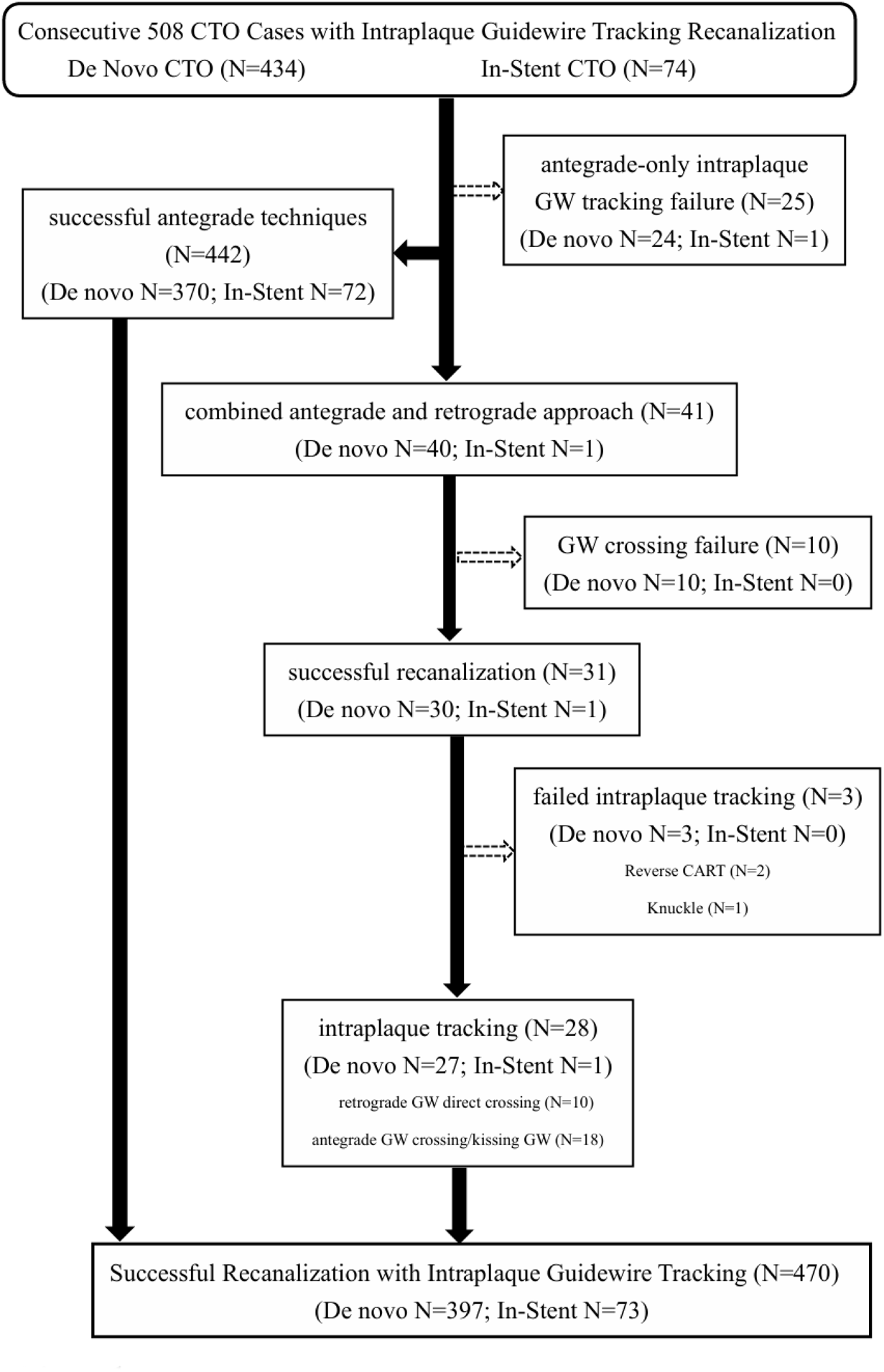
The 508 cases with CTO interventions using wiring-based intraplaque tracking techniques. Successfully wiring was performed in 442 of 467 cases using antegrade-only techniques. Forty-one cases required the back-up of retrograde techniques, and 31 cases had successful recanalization. The two cases with reverse CART and one with knuckle wire techniques were considered failed intraplaque tracking. CTO, chronic total occlusion; CART, controlled antegrade and retrograde tracking; GW, guidewire.

### Procedures of Intraplaque Guidewire Tracking Techniques

By coronary angiography, a CTO lesion was considered if the flow was thrombolysis in myocardial infarction (MI) grade 0 with the duration for at least 3 months.^13,14^ Patients with multivessel disease had stenoses ≥50% in at least two major epicardial coronary arteries. The J-CTO score was based on the consensus of two independent operators (Dr. SC Liu and Dr. CL Lee).

The wiring-based intraplaque tracking strategy used in this study was conceptualized by seven high-volume CTO operators from independent medical centers in northern Taiwan. The purposes were to minimize the extent of subintima creation and stenting and to preserve the antegrade flow for any side branch ≥1.5 mm from 5 mm before to 5 mm after the occluded segment in final coronary angiography.^11^ However, we did not intend to ensure a complete intraplaque course. Additionally, we tried antegrade wiring first for all cases even in the presence of an unfavorable anatomy, such as an ambiguous proximal cap, poor distal vessel quality, or bifurcation at the distal cap. The timing of guidewire escalation/de-escalation and switching to a parallel-wire or retrograde approach was determined by each operator. Once retrograde procedures were necessary, guidewires with softer tip load (mainly ≤1 g) were preferred for direct crossing or serving as the landmark for antegrade guidewire kissing. When the aforementioned techniques for intraplaque tracking failed to cross the occluded segment, reverse controlled antegrade and retrograde tracking or knuckle wire methods were used for rescue. However, both methods, even with successful recanalization, were not considered successful intraplaque tracking to minimize the potential confounding of the current study. The total number of guidewires with different tip loads used in each recanalization procedure was counted. If any of “must preserved” side branches ≥1.5 mm was lost during the procedure even with successful intraplaque tracking, we always attempted to restore the antegrade flow to fulfil our principle. There were no “dissection and reentry” devices used in the study, and intravascular imaging was recommended but not mandatory. Moreover, restoration of thrombolysis in MI grade 3 anterograde flow, postprocedural stenosis of <30%, and the absence of in-hospital major adverse cardiac and/or cerebrovascular events, including cardiac death, Q-wave MI, stroke, or any repeat target lesion revascularization, were fundamental to the definition of procedural success. In patients with successful intraplaque tracking and revascularization, detailed time intervals of guidewire crossing and the total procedure were recorded.

### Statistical Analyses

Statistical analyses were performed using the R 4.3.2 software (R Foundation for Statistical Computing, Vienna, Austria). In statistical testing, two-sided p value ≤ 0.05 was considered statistically significant. The distributional properties of continuous variables are expressed as mean±standard deviations, and categorical variables are expressed as frequencies and percentages. Among the 470 patients with successful recanalization using intraplaque tracking techniques, the comparisons of the guidewire crossing time and the percentage of guidewire crossing time ≥30 min between patients with de novo and in-stent CTOs across the J-CTO scores =0, 1, 2, and ≥3 were analyzed. Subsequently, multivariate logistic regression analysis was performed to calculate the odds ratio (OR) and 95% confidence interval (CI) to select a set of independent features associated with guidewire crossing time ≥30 min among the successfully recanalized CTO lesions with the J-CTO score ≥2. Furthermore, when considering the significantly different percentage of the retrograde approach used between the two groups and its potential effect on guidewire crossing time because of the recanalization principle of the study, we analyzed the effect of J-CTO score on guidewire crossing time in 442 patients with successful antegrade-only procedures (de novo CTO=370, in-stent CTO=72). The differences in the Kaplan–Meier curve of the time for guidewire crossing across each J-CTO score were initially compared using the log-rank test for patients with de novo and in-stent CTOs. Subsequently, the differences in the distributions of continuous and categorical variables across each J-CTO score were examined using the Kruskal–Wallis rank-sum, chi-square, and Fisher’s exact tests as appropriate for the data type. Next, multivariate analysis was performed by fitting a Cox’ proportional hazards model for the time of guidewire crossing to estimate the adjusted hazard ratios of clinical variables and each J-CTO score relative to J-CTO score =0 in patients with de novo and in-stent CTOs. Linear regression analysis of the guidewire crossing time was performed to estimate the adjusted effects of the contributing factors (including the five parameters for counting the total J-CTO score) in patients with de novo and in-stent CTOs. The detailed statistical methods are described in the Appendix.

## RESULTS

### Comparisons of Clinical Features

Among the 508 patients undergoing CTO-PCI in Table 1, those with in-stent CTO (N=74, 14.6%) had higher percentage of diabetes (53 versus 40%, p=0.046), old MI (32 vs. 20%, p=0.017), a history of bypass graft surgery (19 vs. 6%, p<0.001), congestive heart failure (36 vs. 23%, p=0.015), and CKD (32 vs. 23%, p=0.074), compared with those with de novo CTO (N=434). Other clinical features were similar between the two groups.

**Table 1.** Comparisons of clinical features between patients with de novo and in-stent CTO.

|  | **All**<br>**(N=508)** | **de novo**<br>**(N=434)** | **In-stent**<br>**(N=74)** | **P-value** |
| --- | --- | --- | --- | --- |
| Age (yrs) | 64.6±11.6 | 63.8±11.9 | 65.9±9.1 | .146 |
| Male (%) | 446 (88%) | 382 (88%) | 64 (89%) | .693 |
| BMI (kg/m<sup>2</sup>) | 26.2±3.8 | 26.2±3.8 | 26.0±3.6 | .610 |
| Hypertension (%) | 388 (76%) | 332 (76%) | 56 (76%) | .878 |
| Diabetes (%) | 214 (42%) | 175 (40%) | 39 (53%) | .046 |
| Dyslipidemia (%) | 388 (76%) | 327 (75%) | 61 (82%) | .185 |
| Smoking (%) | 180 (35%) | 157 (36%) | 23 (31%) | .398 |
| Old MI (%) | 111 (22%) | 87 (20%) | 24 (32%) | .017 |
| CABG (%) | 39 (8%) | 25 (6%) | 14 (19%) | <.001 |
| CHF (%) | 128 (25%) | 101 (23%) | 27 (36%) | .015 |
| eGFR<60 ml/min/1.73m<sup>2</sup> (%) | 130 (26%) | 104 (23%) | 26 (32%) | .074 |
| Stroke (%) | 32 (6%) | 27 (6%) | 5 (7%) | .861 |

BMI, body mass index; CABG, coronary artery bypass graft; CHF, congestive heart failure; CTO, chronic total occlusion; eGFR, estimated glomerular filtration rate; MI, myocardial infarction

### Comparisons of Angiographic and Procedural Characteristics

Compared with the de novo CTO lesions, the in-stent CTO lesions were located less at the left anterior descending artery (24 vs. 39%, p=0.016) and more at the saphenous venous graft (5% vs. 0, p<0.001) than de novo CTO (Table 2). Combined femoral with radial access as the trans-arterial route for intervention was less frequently used for in-stent than for de novo CTO-PCI (1 vs. 12%, p=0.006). The J-CTO score was lower in patients with in-stent CTO than that in patients with de novo CTO (in-stent: 1.9±1.2 vs. de novo: 2.4±1.4, p=0.005) owing to a lower percentage of blunt stump (in-stent: 27 vs. de novo: 56%, p<0.001), calcification (in-stent: 35 vs. de novo: 58%, p<0.001), and retry cases (in-stent: 5 vs. de novo: 15%, p=0.024). However, an occlusion length ≥2 cm was more frequent in in-stent than in de novo CTO lesions (in-stent: 74 vs. de novo: 52%, p<0.001). Moreover, intravascular imaging (27 vs. 47%, p=0.002), retrograde techniques (1 vs. 9%, p=0.022), parallel wire technique (0 vs. 14%, p=0.001), and antegrade guidewires with ≥3 different tip load (20 vs. 36%, p=0.007) were less commonly used for in-stent than for de novo CTO recanalization. Additionally, shorter procedure and fluoroscopy times, less radiation, and fewer contrast agents were evident in in-stent than in de novo CTO interventions. The success rate of intraplaque guidewire tracking for in-stent CTO was higher than that for de novo CTO (99 vs. 91%, p=0.03). The average time for antegrade wiring before switching to retrograde approach was 62±32 min for all interventions.

**Table 2.** Comparisons of angiographic and procedural characteristics between patients with de novo and in-stent CTO with intraplaque guidewire tracking techniques.

|  | All<br>(N=508) | de novo<br>(N=434) | In-stent<br>(N=74) | P-value |
| --- | --- | --- | --- | --- |
| Multivessel dz. (N) | 458(90%) | 393(91%) | 65(88%) | .470 |
| Lesion Location (N) |  |  |  | . |
| LAD (N) | 187(37%) | 169(39%) | 18(24%) | .016 |
| LCX (N) | 98(19%) | 83(19%) | 15(20%) | .818 |
| RCA (N) | 217(43%) | 180(41%) | 37(50%) | .171 |
| LM (N) | 2(0.4%) | 2(0.5%) | 0 | .559 |
| SVG (N) | 4(0.8%) | 0 | 4(5%) | <.001 |
| All transradial (N) | 374(74%) | 315(72%) | 59(80%) | .174 |
| All transfemoral (N) | 82(16%) | 68(16%) | 14(19%) | .588 |
| Radial+Femoral (N) | 52(10%) | 51(12%) | 1(1%) | .006 |
| J-CTO score | 2.3±1.4 | 2.4±1.4 | 1.9±1.2 | .005 |
| Blunt stump (N) | 262(52%) | 242(56%) | 20(27%) | <.001 |
| Occlusion ≥20mm (N) | 282(56%) | 227(52%) | 55(74%) | <.001 |
| Calcification (N) | 278(55%) | 251(58%) | 27(35%) | <.001 |
| Bending >45°(N) | 296(58%) | 258(59%) | 38(51%) | .192 |
| Retry case (N) | 70(14%) | 66(15%) | 4(5%) | .024 |
| Intravascular Imaging (N) | 227(45%) | 206(47%) | 21(27%) | .002 |
| IVUS (N) | 215(42%) | 197(45%) | 18(24%) | .001 |
| OCT (N) | 12(2%) | 9(2%) | 3(4%) | .379 |
| Multivessel PCI (N) | 212(42%) | 188(43%) | 24(32%) | .080 |
| Retrograde approach (N) | 41(8%) | 40(9%) | 1(1%) | .022 |
| Parallel wire (N) | 59(12%) | 59(14%) | 0 | .001 |
| Antegrade GW≥3 (N) | 173(34%) | 158(36%) | 15(20%) | .007 |
| RA (N) | 18(4%) | 17(4%) | 1(%) | .271 |
| Contrast volume (ml) | 171±82 | 179±85 | 131±49 | <.001 |
| Procedure time (min) | 81±54 | 84±56 | 65±43 | .004 |
| Successful intraplaque tracking (N) | 470 (93%) | 397(91%) | 73(99%) | .030 |
| RAK (Gy) | 5.9±4.7 | 6.1±4.8 | 4.8±4.2 | .041 |
| DAP (mGym<sup>2</sup>) | 37.7±29.9 | 38.9±30.4 | 31.0±25.6 | .053 |
| Fluoroscopy time (min) | 58±34 | 60±35 | 46±29 | .003 |

CTO, chronic total occlusion; DAP, dose area product; GW, guidewire; IVUS, intravascular ultrasound; LAD, left anterior descending; LCX, left circumflex; LM, left main; OCT, optical coherence tomography; PCI, percutaneous coronary intervention; RA, rotational atherectomy; RAK, reference air kerma; RCA, right coronary artery; SVG, saphenous venous graft.

### Application of the J-CTO Score to Procedural Success and Guidewire Crossing Time

The success rate of intraplaque guidewire tracking and crossing for in-stent and de novo CTO interventions are shown in Figure 2. Unlike the significant decrease of success rate for de novo CTOs with the J-CTO score >2 (85 vs. ≤2: 97%, p<0.001), the success rates were comparable between in-stent CTOs with the J-CTO score >2 and ≤2 (96 vs. 100%, p=0.400).

**Figure 2.**
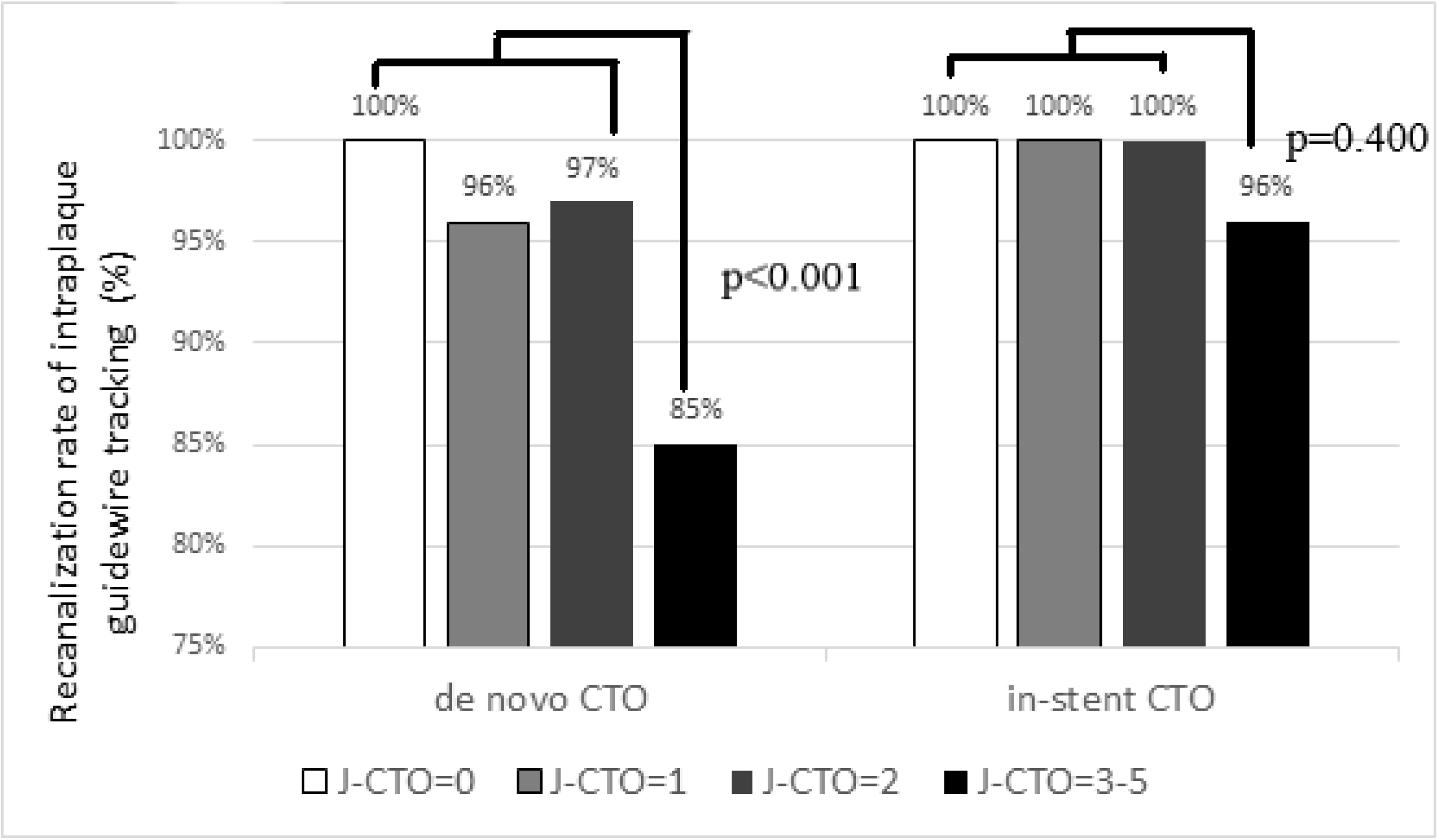
The success rate of intraplaque guidewire tracking for in-stent and de novo CTOs according to the J-CTO Score. CTO, chronic total occlusion.

Among the 470 patients with successful recanalization using intraplaque guidewire tracking techniques, the mean guidewire crossing time was significantly longer for patients with de novo CTO (397 patients) than that for those with in-stent CTO (73 patients) (28±35 vs. 16±22 min, p=0.005). Although the comparisons of guidewire crossing time between the de novo and in-stent CTOs across each J-CTO score did not reach statistical significance (Figure 3A), the difference in the percentage of guidewire crossing time ≥30 min between the two groups was more evident when the J-CTO score was ≥3 (de novo: 51% vs. in-stent CTO: 28%, p=0.03) (Figure 3B). Among those with the J-CTO score ≥2, there was a significantly lower chance of guidewire crossing time ≥30 minutes for in-stent than for de novo CTO recanalization (OR=0.40, 95% CI=0.18–0.86) in multivariate analysis after correcting for age, sex, and presence of multivessel disease.

**Figure 3.**
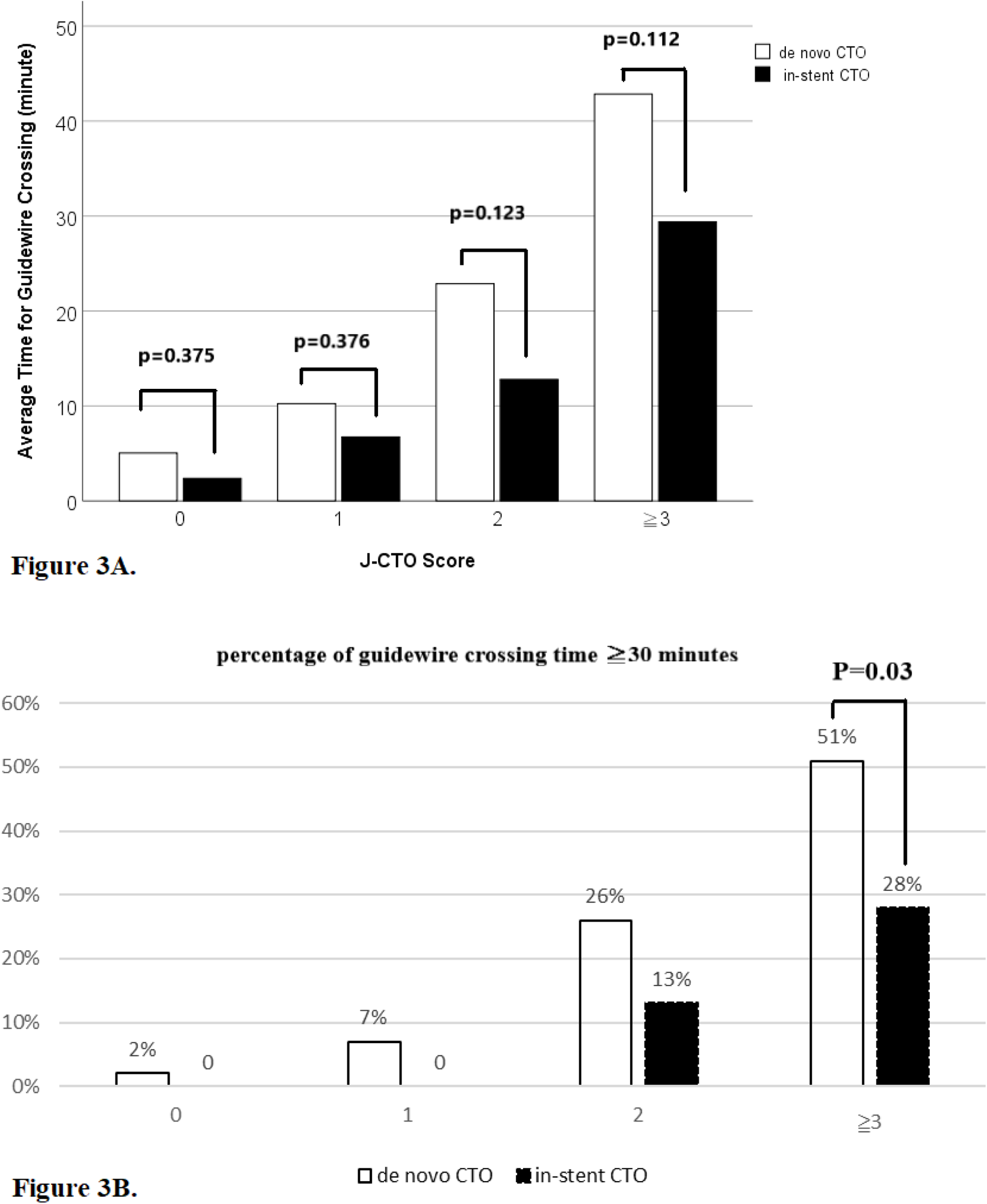
Comparisons of the guidewire crossing time (Figure 3A) and the percentage of guidewire crossing time ≥30 minutes (Figure 3B) with respect to each J-CTO Score between patients with in-stent versus de novo CTO lesions recanalized successfully by intraplaque guidewire tracking. CTO, chronic total occlusion.

### Effects of Clinical Features and Each J-CTO Score on Guidewire Crossing Time in 442 Patients with Successful Recanalization using Antegrade-Only Intraplaque Guidewire Tracking Techniques

The differences in the Kaplan–Meier curve of the time for guidewire crossing across each J-CTO score were significant for 370 patients with de novo CTO (p<0.001 by log-rank test, Figure 4A) and 72 patients with in-stent CTO (p<0.001 by log-rank test, Figure 4B) who were successfully revascularized via antegrade-only intraplaque wiring-based tracking techniques. In the multivariate Cox’ proportional hazards model, the decreasing adjusted hazard ratios of guidewire crossing time with increasing J-CTO score relative to J-CTO=0 were significant for both the de novo and in-stent CTO groups (Table 3).

**Figure 4.**
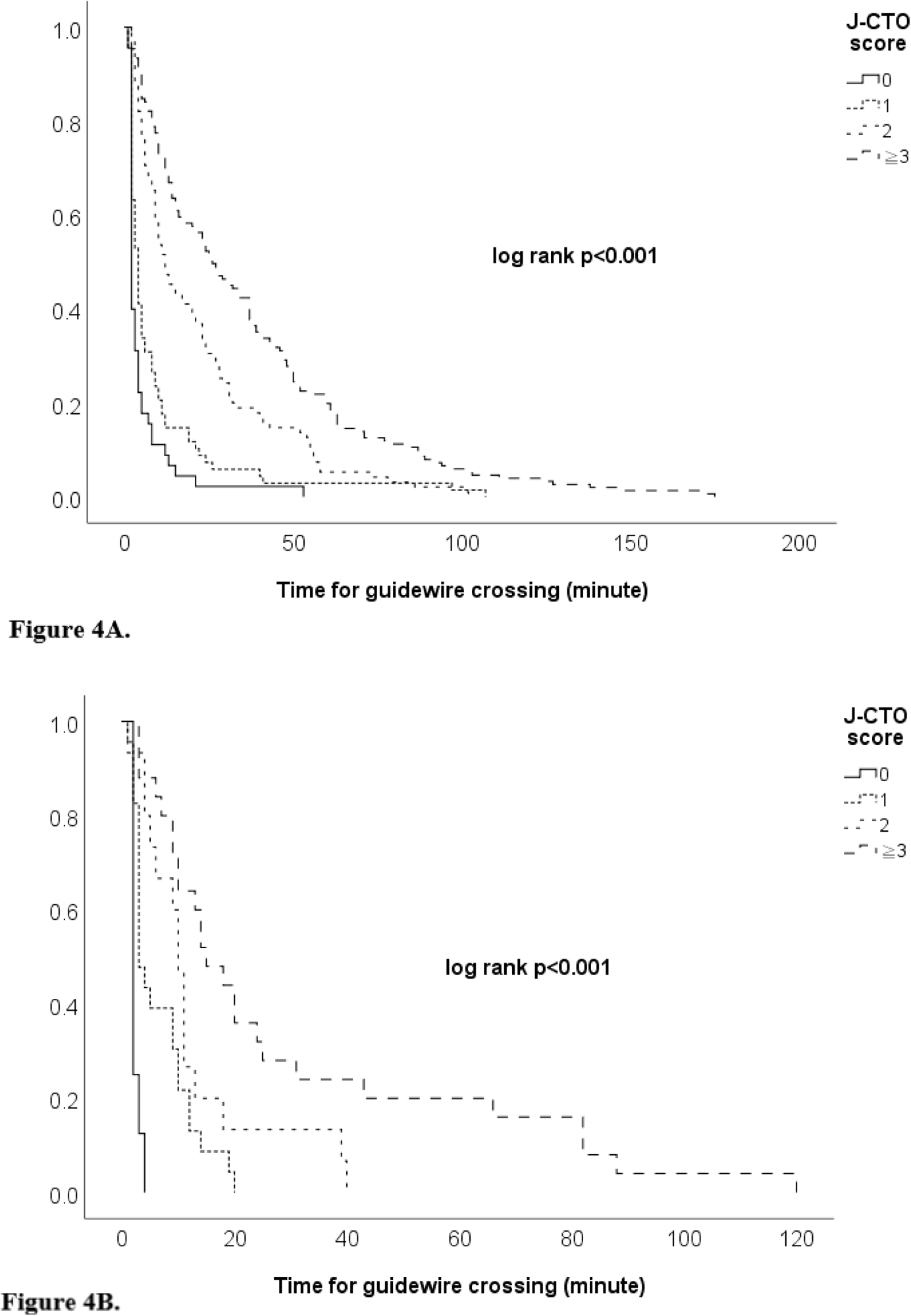
The Kaplan–Meier Curves showed the comparisons of guidewire crossing time with respect to each J-CTO score for patients with de novo (Figure 4A) and in-stent (Figure 4B) CTOs recanalized successfully by antegrade-only intraplaque guidewire tracking techniques. CTO, chronic total occlusion.

**Table 3.**
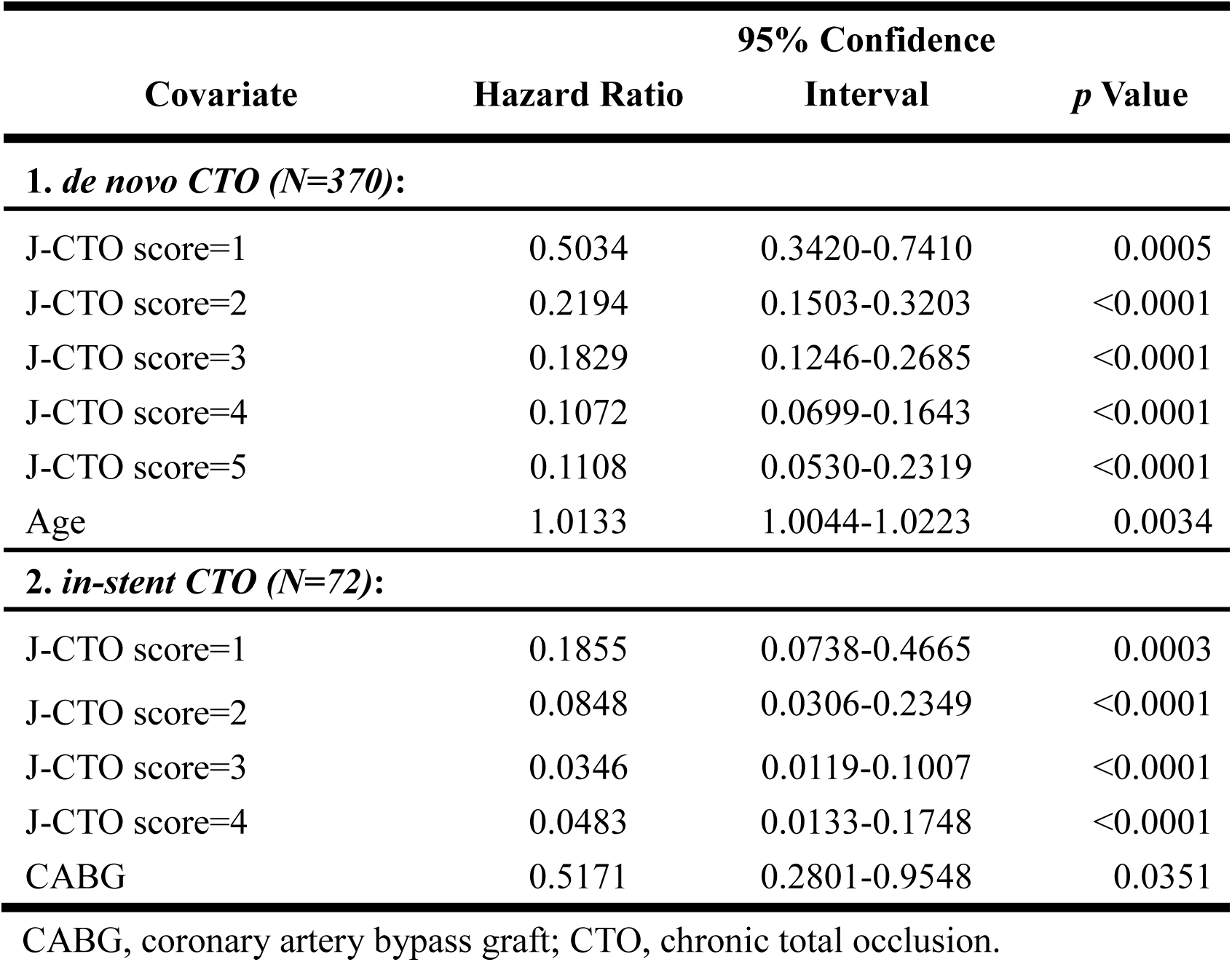
Cox’s proportional hazards model for the time of guidewire crossing to estimate the adjusted hazard ratios of clinical features and each J-CTO score relative to J-CTO score = 0 in 442 patients with de novo CTO and in-stent CTO successfully recanalized by antegrade intraplaque guidewire tracking techniques, respectively.

### Contributions of Each Parameter for Counting J-CTO Score to Guidewire Crossing Time in 442 Patients with Successful Recanalization using Antegrade-Only Intraplaque Guidewire Tracking Techniques

In multiple linear regression analysis (Table 4), the guidewire crossing times were independently increased by 7.6±3.0 min, 14.0±3.0 min, 12.8±3.0 min, and 10.8±4.3 min (all p<0.02) with the presence of blunt stump, occlusion ≥20mm, bending >45, and retry procedure, respectively, for 370 patients with de novo CTO recanalized via antegrade-only intraplaque guidewire tracking techniques. However, for 72 patients with in-stent CTO, the time of guidewire crossing was increased only by the presence of blunt stump and occlusion ≥20mm for 15.0±5.6 min and 16.2±5.6 min, respectively (both p<0.01).

**Table 4.** Multiple linear regression analysis of the guidewire crossing time to estimate the adjusted effects of the contributing factors (including the five parameters for counting the total J-CTO score) in 442 patients with de novo CTO and in-stent CTO recanalized successfully by antegrade intraplaque guidewire tracking techniques, respectively.

| Covariate | Parameter Estimate | Standard Error | *t* Value | Pr > \|*t*\| |
| --- | --- | --- | --- | --- |
| **1. De novo CTO (N=370):** |  |  |  |  |
| Intercept | 4.8886 | 2.3872 | 2.0479 | 0.0413 |
| Blunt stump | 7.6293 | 2.9513 | 2.5851 | 0.0101 |
| Occlusion $\geq 20$mm | 13.9718 | 2.9731 | 4.6994 | <0.0001 |
| Bending >45° | 12.8015 | 3.0286 | 4.2268 | <0.0001 |
| Retry | 10.8485 | 4.2997 | 2.5231 | 0.0121 |
| **2. In-stent CTO (N=72):** |  |  |  |  |
| Intercept | -0.3561 | 5.0586 | -0.0704 | 0.9441 |
| Blunt stump | 14.9611 | 5.5560 | 2.6928 | 0.0089 |
| Occlusion $\geq 20$mm | 16.1928 | 5.5560 | 2.9145 | 0.0048 |

CTO, chronic total occlusion.

## DISCUSSION

The percentage of in-stent CTO in the registry (14.6%) is comparable with that (14.7%) of the pooled data of four multicenter registries.^3^ The main findings of the study are as follows: (1) With intraplaque guidewire tracking techniques for in-stent CTO recanalization, the J-CTO score, unlike that for de novo CTO, failed to predict procedural success between the very difficult (>2) and the relatively easier (≤2) lesions. (2) There was less need of guidewire crossing time ≥30 min for in-stent than for de novo CTO with the J-CTO score ≥2 among those with successful recanalization. (3) The time of guidewire crossing by antegrade-only techniques for recanalizing in-stent CTO was still relevant to the J-CTO score. However, only a blunt stump and occlusion ≥20mm served as determining factors for the increased time for guidewire crossing. These findings suggest that the J-CTO score should be interpreted with caution when evaluating interventions for in-stent CTO.

Several studies have validated the increased risk of recanalization failure with a higher J-CTO lesion complexity score.^15–18^ When comparing patients with a J-CTO score ≥3 versus <3, a 2.2-fold risk of procedural failure was also shown.^15^ Although some of these studies included a variable percentage up to 16% of in-stent occlusions for analysis,^15,17,18^ the true effect of J-CTO score on recanalization difficulty for in-stent CTO remains uncertain. The current study demonstrated that a significant decline of procedural success rate for de novo CTO interventions with the J-CTO score ≥3 was not evident for in-stent CTO. There are several possible reasons for this observation. First, the significantly higher overall success rate (99%) of in-stent CTO interventions could have led to an undifferentiated effect of the J-CTO score on the difficulty of recanalization. This can be partly because of a numerically lower J-CTO score of 1.9±1.2 for the in-stent group in the present study, compared with that of 2.32±1.26 with a procedural success rate of 85% in the pooled registries.^3^ Additionally, a higher rate of procedural failure has been described in several CTO studies addressing patients with bypass surgery.^19–21^ Therefore, a lower percentage of 19% with prior bypass graft of the in-stent CTO group in our registry than that of 26.5% in the pooled analysis^3^ can be another contributory factor of the higher success rate. Second, the gap in success rates, which was theoretically related to the differences in J-CTO scores between in-stent and de novo CTO interventions, could possibly lead to diverse study results. For example, the pooled data of four registries included an in-stent CTO group with a numerically close but statistically higher J-CTO score than the de novo CTO group (2.32±1.26 vs. 2.22±1.27, p=0.0111), and showed a similar rate of procedural success between the two groups (84.9 vs. 85.2%, p=0.75).^3^ Lee et al. analyzed another multicenter registry that showed that even if the in-stent CTO group had a significantly higher J-CTO score than the de novo CTO group (2.07±1.21 vs. 1.68±1.27, p<0.001), there was still a remarkably higher success rate in the in-stent than in the de novo CTO group (84.6 vs. 76.0%, p=0.035).^10^ If we considered the effect of the J-CTO score on recanalization difficulty for de novo CTO as the reference, all studies, including ours (in-stent: 1.9±1.2 with success rate:99% vs. de novo: 2.4±1.4 with success rate: 91%, p<0.05 for comparisons of J-CTO score and success rate), suggested that the association between increasing J-CTO score and the risk of recanalization failure for in-stent CTO could be not identical to that for de novo CTO. Additionally, the difference in the success rate between the two groups was partly because of the techniques used for recanalization. The multicenter registry suggested a technique preference of antegrade wiring for in-stent than for de novo CTO interventions to achieve a similar success rate.^3^ As we used the same strategy as antegrade-first intraplaque guidewire tracking techniques for all patients in our registry, its favorable effect on in-stent CTO recanalization can contribute to the significant difference in the success rate and statistical results between the two groups.

Therefore, we further demonstrated the different effects of J-CTO scores on in-stent versus de novo CTO interventions by analyzing the time for guidewire crossing. Based on the original J-CTO score study for de novo CTO lesions, the chance of guidewire crossing time ≥30 min was considered to be >50% if the score was ≥2.^4^ In the current study, even for difficult and very difficult (J-CTO≥2) in-stent occlusions, there was a 60% of reduced risk for guidewire crossing time ≥30 min compared with that for de novo occlusions in multivariate analysis. For intraplaque guidewire tracking techniques, an in-stent CTO can be easier to cross than a de novo CTO because the visible stent serves as the roadmap, particularly when the occluded segment is longer with unclear tortuosity. Clear guidance would facilitate faster wiring and possibly shorten the time required for a more precise judgement of guidewire escalation. Therefore, a relatively shorter time for guidewire crossing for each J-CTO score (Figure 3A) and a lower need for antegrade guidewires with three different tip loads (20% vs. de novo: 36%, p=0.007) for in-stent CTO recanalization were also suggested in this study.

Although the differential effect of each J-CTO score category on the time of guidewire crossing for in-stent versus de novo CTO recanalization was evident, we used the Kaplan–Meier curves and included patients with successful antegrade-only procedures to show that the J-CTO score was still clearly relevant to the guidewire crossing time for in-stent or de novo CTO recanalization. The positive correlation between the J-CTO score and time of CTO interventions is mostly described as fluoroscopic or procedural time.^16,22,23^ Theoretically, the J-CTO score, which is representative of lesion complexity, must be linked directly to the time of guidewire crossing. Nombela-Franco et al. analyzed 209 patients with CTO-PCI and showed that the guidewire working times were 8, 24, 30, and 69 min for lesions with J-CTO scores of 0, 1, 2, and ≥3, respectively.23 However, the percentage of in-stent occlusions was not described in the study, and retrograde approach was notably used for 53.1% of all recanalization procedures. As the time of crossing collateral channels is irrelevant to lesion complexity or the J-CTO score, counting it in the total guidewire working time would cause potential heterogeneity when studying their relationship. In the present study, we analyzed those with successful recanalization using antegrade intraplaque guidewire tracking and demonstrated that when taking the time of guidewire crossing for lesions with J-CTO=0 as a reference, there was a significantly reduction in the possibility of guidewire crossing with an increasing J-CTO score for either in-stent or de novo occlusions in the multivariate Cox’ proportional hazards model (Table 3).

We further identified that with antegrade intraplaque tracking techniques, each parameter, except calcification of the J-CTO score independently contributed to the time of successful guidewire crossing for de novo CTO recanalization. However, only the presence of a blunt stump and an occluded length ≥20 mm were relevant factors for in-stent occlusion. In our previous study, we suggested that the presence of calcification did not prolong the time of guidewire crossing using intraplaque guidewire tracking techniques for CTO-PCI.^24^ The reason could be that the distribution of calcification serving as the roadmap would partly facilitate guidewire coursing through the occluded route. Additionally, when calculating the J-CTO score of in-stent CTO, the definition of calcification outside or inside the stent has not yet been clearly described. If defined as outside the stent, some calcification can be more easily obscured by the stent struts and judged as non-calcification for in-stent than for de novo CTO lesions. However, calcification inside the stent, mostly due to neoatherosclerosis, cannot be a likely definition because the rates of intravascular imaging use were <40% in most in-stent CTO studies.^3,6,8,9^ If calcium is outside the stent, its effect on wiring procedures can be significantly less than that for de novo CTO interventions. Furthermore, bending >45°, with procedural significance for de novo CTO recanalization, was not a time-determining factor for guidewire crossing for in-stent CTO in the current study. Similarly, the stent itself delineates the clear curvature of the occlusion and significantly increases the likelihood of intraplaque guidewire tracking even in lesions with high angulation. This can lead to a diminished effect of high angulation on the time of guidewire crossing for in-stent than for de novo CTOs. With respect to the limited role of previous failure attempt in the guidewire crossing time of in-stent CTO-PCI shown in the study, the interpretation should be more conservative because of the significantly fewer cases (4 cases, 5%) in our registry and deserve further clarification.

### Study Limitations

The study has some limitations. First, the study results could have been altered if the revascularization strategy had changed. For example, if the use of dissection/reentry devices or techniques with either antegrade or retrograde strategies is favored for CTO recanalization, the effect of each J-CTO parameter on these procedures must differ from that on intraplaque wiring. However, this study aimed to compare the application of the J-CTO score for in-stent and de novo CTO interventions. The same wiring-based CTO recanalization strategy would minimize potential heterogeneity if multiple strategies were involved. Second, intraplaque tracking and crossing were not completely confirmed using intravascular imaging. As described in the Methods section, the principle of intraplaque tracking is to try to minimize subintima creation and stenting, but not to ensure a 100% intraplaque course. One CTO-PCI study suggested that the discordance between presumed and intravascular ultrasound-confirmed true lumen was only 15.8%.^25^ Moreover, the definition of procedural success in the study requiring preservation of all significant side branches would further suggest the limited subintima creation.

## CONCLUSIONS

With intraplaque guidewire tracking techniques, this study suggested that the application of the J-CTO score to in-stent versus de novo CTO interventions was different with respect to its effects on the rate of procedural success for lesions with higher complexity, the probability of guidewire crossing time ≥30 min when the score was ≥2. Although the increasing J-CTO score is associated with more prolongation of guidewire crossing time, only the presence of a blunt stump and an occluded length ≧20 mm independently contributed to the time for guidewire crossing. Therefore, whether any lesion factors, in addition to the J-CTO score, can serve to evaluate the recanalization difficulty of in-stent occlusions deserves further investigation and clarification.

## Data Availability

The data that support the findings of this study are available from the corresponding author upon reasonable request.

## ACKNOWLEDGMENTS

Grant from the Taiwan Health Foundation, Taipei, Taiwan.

